# Meta-analysis of several epidemic characteristics of COVID-19

**DOI:** 10.1101/2020.05.31.20118448

**Authors:** Panpan Zhang, Tiandong Wang, Sharon X. Xie

## Abstract

As the COVID-19 pandemic has strongly disrupted people’s daily work and life, a great amount of scientific research has been conducted to understand the key characteristics of this new epidemic. In this manuscript, we focus on four crucial epidemic metrics with regard to the COVID-19, namely the basic reproduction number, the incubation period, the serial interval and the epidemic doubling time. We collect relevant studies based on the COVID-19 data in China and conduct a meta-analysis to obtain pooled estimates on the four metrics. From the summary results, we conclude that the COVID-19 has stronger transmissibility than SARS, implying that stringent public health strategies are necessary.

## 1 Introduction

Coronavirus disease (COVID-19) is an infectious disease caused by *severe acute respiratory syndrome coronavirus 2* (SARS-CoV-2), a newly discovered coronavirus, which leads to respiratory illness and can be transmitted from person to person. Ever since December 2019, when the first case of COVID-19 in Wuhan, P. R. China (or mainland China) was reported, the novel coronavirus has hit most of the countries in the world, with the United States (U.S.) being the one having the largest number of confirmed cases (Worldometer, 2020). On March 11, 2020, the World Health Organization (WHO) declared COVID-19 a worldwide pandemic. As of May 9, 2020, the WHO has reported a total of 3,855,788 confirmed cases all over the world and the total number of deaths has reached 265,862.

The COVID-19 pandemic has significant negative impacts on both the global health and the economy. In the U.S., for example, the unemployment rate has jumped up to 14.7% in April, 2020, reaching the highest rate and the largest monthly increase since January 1948 (U.S. Bureau of Labor Statistics, https://www.bls.gov/bls/newsrels.htm). Continuous efforts have been made by every country in the world to slow the spread of the disease and mitigate the associated negative impacts on various aspects of the society.

So far, great amount of scientific research has been conducted on COVID-19, ranging from ongoing clinical trials that evaluate potential treatments to statistical analyses on the characteristics of this infectious disease. With more and more COVID-19 data and studies available, it is vitally important to aggregate the information and pool the statistical findings. Hence, here we conduct a meta-analysis based on published studies of the COVID-19 outbreak in mainland China.

Our meta-analysis that accounts for between-study heterogeneity concentrates on four common epidemic metrics that characterize the transmission of the COVID-19:

1. *Basic reproduction number*: Often referred to as *R*_0_, the basic reproduction number measures the contagiousness or transmissibility of infectious agents and is interpreted as the expected number of infections caused directly by one case in a completely susceptible population.
2. *Incubation period*: This metric is defined as the number of days between when an individual was actually infected and when this person starts to show symptoms. Understanding the incubation period of the COVID-19 is crucial as it provides guidelines on deciding a reasonable length of the quarantine period.
3. *Serial interval*: Defined as the time between the start of symptoms in the primary patient (infector) and onset of symptoms in the patient being infected by the infector (the infectee), the serial interval is critical in the calculation of *R*_0_.
4. *Epidemic doubling time*: This metric measures the period of time needed for the total number of cases in the epidemic to double, and is an important factor that reflects the speed at which the COVID-19 is spreading.

The rest of this manuscript is organized as follows. We first provide details on the data collection process in Section 2. Then in Section 3, we introduce the key ingredients for a meta-analysis, where the modeling and estimation methods are outlined. Section 4 lists a detailed summary for the four epidemic metrics as given above. Statistical results from the meta-analysis are reported in Section 5, and sensitivity analysis is given in Section 6. Concluding remarks are then disclosed in Section 7.

## 2 Study selection

We conduct a comprehensive literature screening for the articles published on scientific journals (including early versions) or preprint platforms like *medRxiv* and *bioRxiv*. The key words of our searching are COVID-19 (SARSCoV-2 or 2019-nCOV), the epidemic characteristics of interest (basic reproduction number, incubation period, serial interval and epidemic doubling time), and China, where the selection criteria is relaxed for regional labels. Specifically, we include the studies on the whole country of China, China except for Hubei province (when the city of Wuhan is located), a list of provinces and municipalities, a collection of cities or even a specific region, in order to increase the sample size and the potential statistical power. The variability and heterogeneity across the selected studies are accounted when adopting appropriate methodologies in the meta-analysis.

In addition, we only include the studies reporting unambiguous estimates and the associated 95% confidence intervals or standard deviations. If the standard deviation of an estimator is given, we calculate the 95% confidence interval under the assumption of normal approximation provided that the fitted model is valid. This selection criterion is needed to ensure the consistency of analysis results.

Some studies included here provide more than one estimates that are obtained from various methods or models. For those studies, we select one estimate and its associated confidence interval in our analysis. We illustrate our choices and the corresponding reasons in details in Section 5.

## 3 Meta-analysis

*Meta-analysis* is a statistical procedure aiming to combine scientific results from multiple comparable studies or trials. It is one of the most popular analytical tools in the statistical analysis, which derives a pooled estimate by aggregating relevant information, thus increasing the statistical power. In particular, when comparing different studies addressing the same question, the crux is to measure the standardized difference across various results, i.e. the *effect size*. In this sense, we rely on meta-analysis to improve the estimates of the effect size of an intervention or an association and examine variability to detect inconsistent results across studies.

In general, there are two methods to pool effect sizes from multiple studies: the *fixed-effects model* (FEM) and the *random-effects model* (REM). A FEM assumes all included studies come from the same population, whereas a REM is constructed based on the assumption that data collected comes from different population. Although we restrict our searching of COVID-19 research with data only from mainland China, difference in the population may still exist, due to possible variations in the data samples as well as sampling errors. Hence, we choose to adopt the REM in our analysis.

Let *k* index the selected studies and *Y_k_* be the estimator of the *k*-th study. A REM assumes a *normal-normal hierarchical* model:

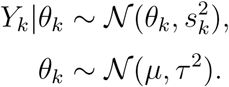

Here *θ_k_* is the parameter of interest, and 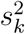 is the variance of *Y_k_*. The hyper-parameters *µ* and *τ*^2^correspond to the true effect and the across-study variance that reflects heterogeneity of the population, respectively.

The inference for REM is done sequentially: we first estimate the heterogeneity variance *τ*^2^(denoted as 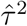), then with 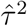 given, we obtain estimates of the effect *µ*. There are a variety of estimation methods for *τ*^2^; see Veroniki et al. (2016) for a concise survey. We adopt the *Hartung-Knapp-Sidik-Jonkman* (HKSJ) method (Hartung and Knapp, 2001; Sidik and Jonkman, 2002), which is shown to be more robust when the number of studies is small and there is a substantial heterogeneity in the population.

Given 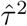, the conditional maximum-likelihood of effect estimate becomes

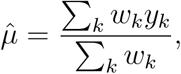

where the weights are specified as

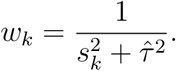

Then we use the normal approximation to construct confidence intervals, where the standard deviation of 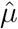 is given by

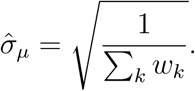

Later in Section 5, we also provide prediction intervals, which are equally significant components in a meta-analysis. In the presence of a substantial across-study heterogeneity, prediction intervals are preferred as they not only quantitatively provide a range for the effect size of a new study, but also measure the uncertainty of the estimate in a way that acknowledges the heterogeneity.

## 4 Epidemic characteristics

In this section, we briefly introduce the epidemic characteristics that are investigated in this manuscript.

### Basic reproduction number

In epidemiology, the *basic reproduction number*, denoted *R*_0_, is the expected number of cases caused by one case in a completely susceptible population. It is a critical metric to describe the contagiousness or transmissibility of infectious diseases (Delamater et al., 2019). The estimation of *R*_0_ is primarily based on compartmental models, where the *susceptible-infectious-recovered* (SIR) model and its extensions are the most commonly used. A variety of methods have been developed to estimate *R*_0_, such as maximum likelihood methods and Bayesian approaches; we refer the interested readers to Dietz (1993) for a concise survey and to Nikbakht et al. (2019) for a practical comparison of the methods. For the users of statistical software R, the package R0 includes most standard methods for the estimation of *R*_0_.

Since the outbreak of COVID-19, *R*_0_ has been one of the most critical metrics receiving substantial interest in the community, as it provides a basic benchmark (with threshold 1) to define a pandemic. Besides, *R*_0_ helps indicating the potential severity of an epidemic outbreak. It is evident that *R*_0_ is closely related to the fraction of the number of infected individuals out of a population once the outbreak ends (Holme and Masuda, 2015). Furthermore, *R*_0_ is an indispensable component when estimating the effective reproduction numbers (denoted as *R_t_*), which are often used to assess the effectiveness of the intervention procedure to mitigate the spread of an epidemic.

### Incubation period

The second characteristic that we investigate here is the incubation period. The incubation period of an epidemic is the period from the time of the contact of a transmission source (susceptible or confirmed infector) and the time of symptom onset. The incubation period provides important information during an outbreak, as it helps to determine when the infected individuals who are symptomatic are most likely to spread the disease, and signal necessary public health activities such as monitoring, surveillance and intervention.

From a statistical perspective, the incubation period is a crucial factor to model the current and future trends of an epidemic, as well as evaluate intervention strategies. One of the biggest challenges of estimating the incubation period is that data are often coarsely observed, in addition to several other issues such as censoring and selection bias. The estimation of incubation period is generally based on the methods developed in Reich et al. (2009) and the extensions.

### Serial interval

The *serial interval* is defined as the time duration between the onset of symptoms in the primary patient and the onset of symptoms in the secondary patient who receives the disease from that primary one (Lipsitch et al., 2003). It is the sum of the latent period and the duration of infectiousness. Being another crucial factor for constructing epidemiological models, the serial interval is one of the fundamentals for computing and estimating *R*_0_; see Fine (2003) for a summary of the importance of serial interval in epidemiological studies.

The estimation of serial interval is based on the generation time distribution, which is closely related to the infection rate of a epidemic. Standard estimation procedures are given in Diekmann et al. (2013, Chapter 13). In practice, the `R` package `R0` collects functions that estimate serial intervals via maximum likelihood methods (White et al., 2009) and the estimation of serial interval in the package `EpiEstim` is based on the method developed in Cori et al. (2013).

### Epidemic doubling time

The *epidemic doubling time* (or simply doubling time) is another important index in epidemic studies, as it measures the length of the period during which the number of confirmed cases is doubled. It is evident that the doubling time is reversely related to another epidemic parameters of interest: case-fatality rate. Hence, learning doubling time helps epidemiologists not only understand the transmissiblity of an infection disease but also evaluate its severity. In the course of pathology, the doubling time is useful for analyzing the growth rate of viruses and antigens. Doubling time also can be used to assess the effectiveness of public health interventions and protocols, as an increase in the doubling time usually indicates a slowdown in epidemic transmission.

The estimation of the epidemic doubling time is generally model based, where exponential growth models are the most frequently adopted (Galvani et al., 2003; Du et al., 2020b). In this manuscript, we conduct a meta-analysis on the doubling time to gauge the growth rate of COVID-19.

## 5 Results

In this section, we apply the methods introduced in Section 3 to estimate the epidemic characteristics of interest listed in Section 4. The analysis is primarily done in `R`, where several standard packages for meta-analysis are utilized; for instance, `meta`, `metafor`, `dmetar` among others.

### Basic reproduction number

Relevant studies used for our meta-analysis are summarized in Table 2, where a total of 12 research articles are included. Note that the sample size is not large, so we depict a *funnel plot* in Figure 1 to see whether there is any bias owing to the small sample size. In a funnel plot, a significantly asymmetry pattern indicates publication bias. A more rigorous method is to exploit the *Egger’s test* (Egger et al., 1997), for which a small *p*-value suggests rejecting the null hypothesis (i.e., some bias caused by the small sample size does exist). The *p*-value here is 0.47, showing that no further procedure is needed for correcting the effect size for the present meta-analysis.

**Figure 1:**
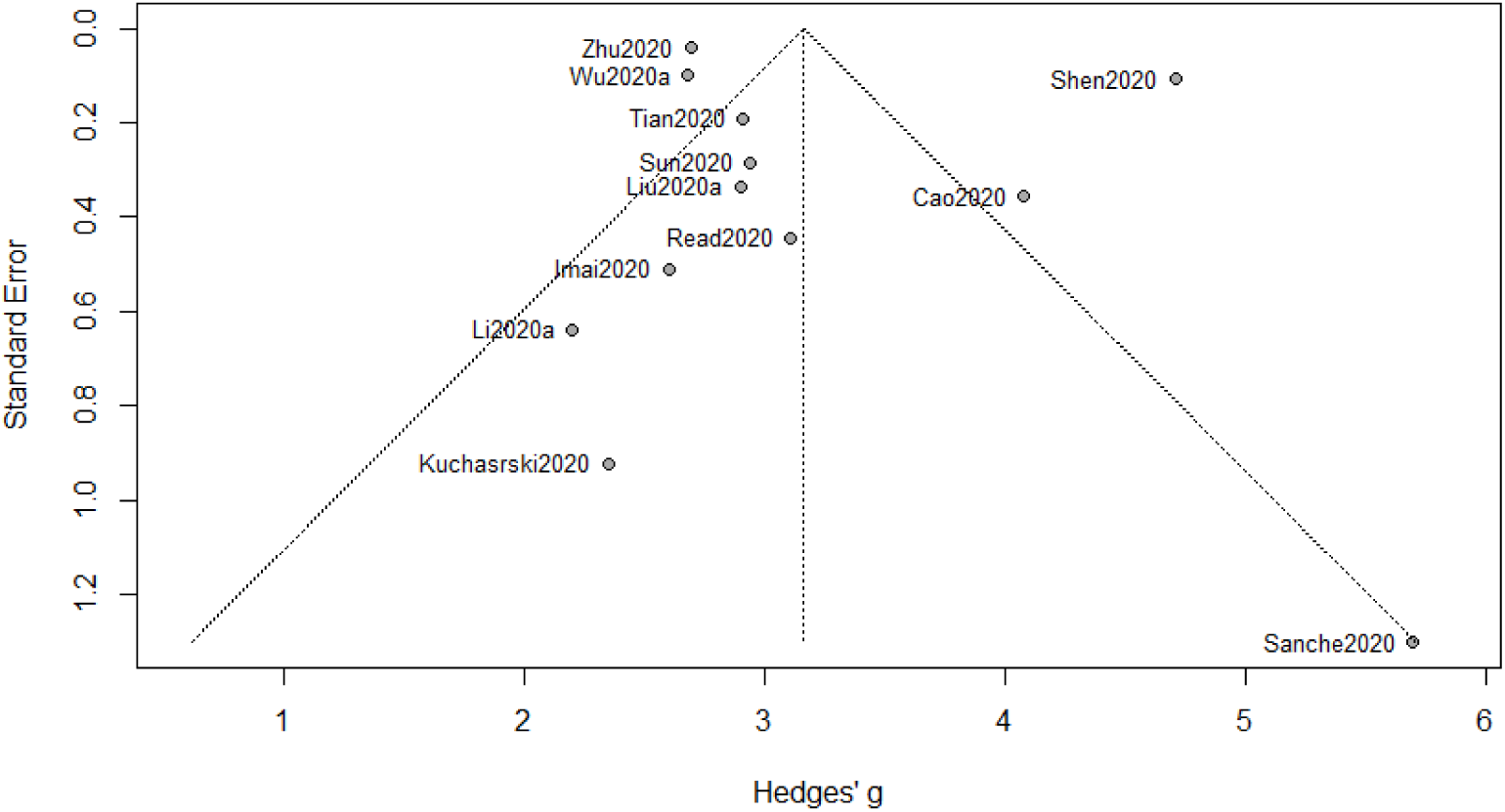
Funnel plot of meta-analysis for *R*_0_

Using the estimation procedure demonstrated in Section 3, we see that the estimate of *R*_0_ is 3.16 with a 95% confidence interval [2.60, 3.72] and a 95% prediction interval [1.25, 5.08]. The associated *forest plot* is presented in Figure 2. The index of heterogeneity, *I*^2^, which ranges from 0 to 1, is used to quantify the dispersion of effect sizes. Here we have *I*^2^ = 97%, indicating a substantial heterogeneity in the population. The conclusion from *I*^2^ is also consistent with a small *p*-value from the *Cochran’s Q* test (less than 10^*−*4^). Hence, the REM is indeed more appropriate than the FEM for our meta-analysis.

**Figure 2:**
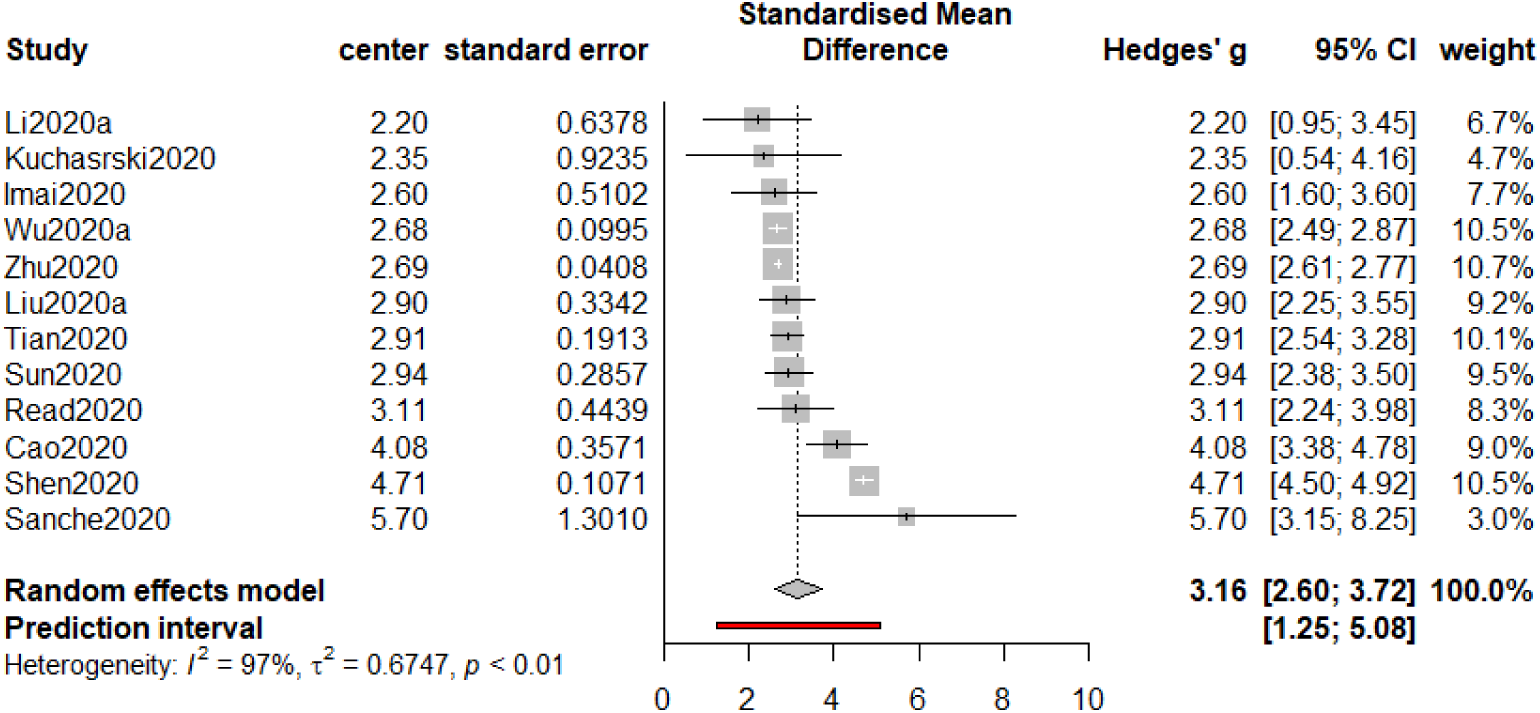
Forest plot of meta-analysis for *R*_0_

With high heterogeneity in the underlying population, prediction intervals that incorporates heterogeneity is more informative than confidence intervals which focuses only on summary estimates. We hence recommend using the prediction interval as an interval estimator when inferring *R*_0_.

Based on the results from our meta-analysis, we conclude that the basic reproduction number (*R*_0_) of COVID-19 appears to be greater than that of SARS (point estimate around 3), as reported by WHO in “Consensus document on the epidemiology of severe acute respiratory syndrome (SARS)”. However, it is not as large as an average-based estimate (3.28) for COVID-19 reported in Liu et al. (2020b). Compared with the results from other two meta-analyses on COVID-19, our estimate is slightly greater than 3.15 reported in He et al. (2020a), and moderately greater than 3.05 reported in Dong et al. (2020). Nonetheless, we do not observe statistically significant difference in either case.

### Incubation period

Analogous to the previous part, we give the funnel plot in Figure 3, from which a (roughly) symmetric pattern is observed. This is consistent with the *p*-value (0.915) from the Egger’s test. Hence, again, publication bias is not present here.

**Figure 3:**
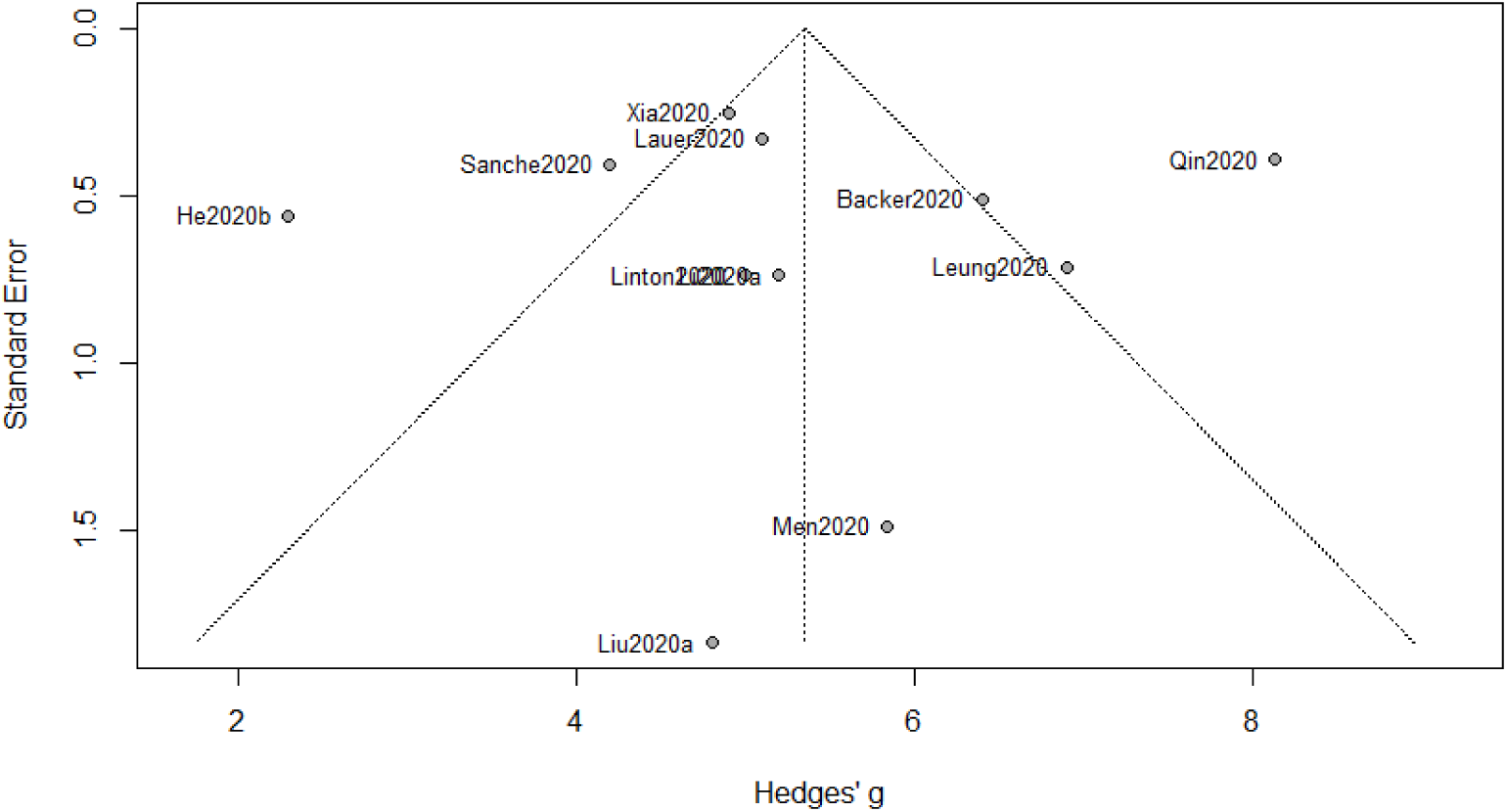
Funnel plot of meta-analysis for incubation period

The estimate of the incubation period of COVID-19 based on our meta analysis is 5.35 (days), with a 95% confidence interval [4.29, 6.42] and a 95% prediction interval [1.97, 8.73]; see Figure 4. Our result is greater than the median (4 with *interquartile range* (IQR) from 2 to 7) of the incubation period estimated by Guan et al. (2020), which is not included in our meta-analysis as no 95% confidence interval is provided therein. One possible reason is that the study period of Guan et al. (2020) is between December 11, 2019 and January 29, 2020, which is considered as a relatively early stage of the COVID-19 outbreak in mainland China. Besides, our estimate is larger than that (5.08) from another meta-analysis in He et al. (2020a).

**Figure 4:**
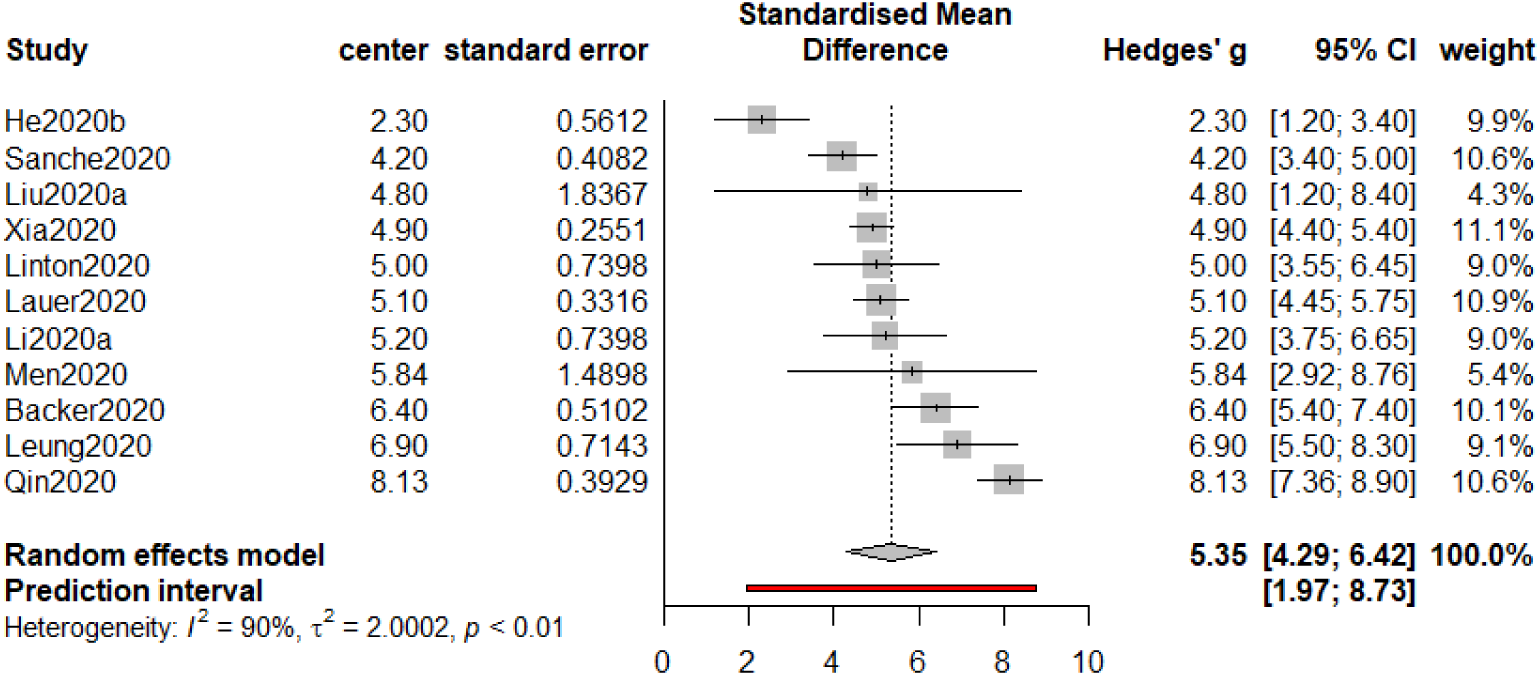
Forest plot of meta-analysis for incubation period

Our meta-analysis also suggests that the incubation period of COVID-19 is a bit longer than that of SARS (commonly ranging from 3 to 5 according to WHO), and our finding is close to the estimate of the incubation period of COVID-19 by the U.S. Centers for Disease Control and Prevention (CDC). Our result provides evidence in support of the 14-day monitoring and quarantine periods in implementation.

### Serial interval

For this part, we collect 12 studies to proceed along our meta-analysis. Albeit the funnel plot in Figure 5 displaying an asymmetric pattern, the *p*-value from the Egger’s test is 0.29, suggesting that it is not required to implement a correction procedure. The estimate of the serial interval is 5.35 with a 95% confidence interval [4.63, 6.07] and a 95% prediction interval [3.13, 7.57].

**Figure 5:**
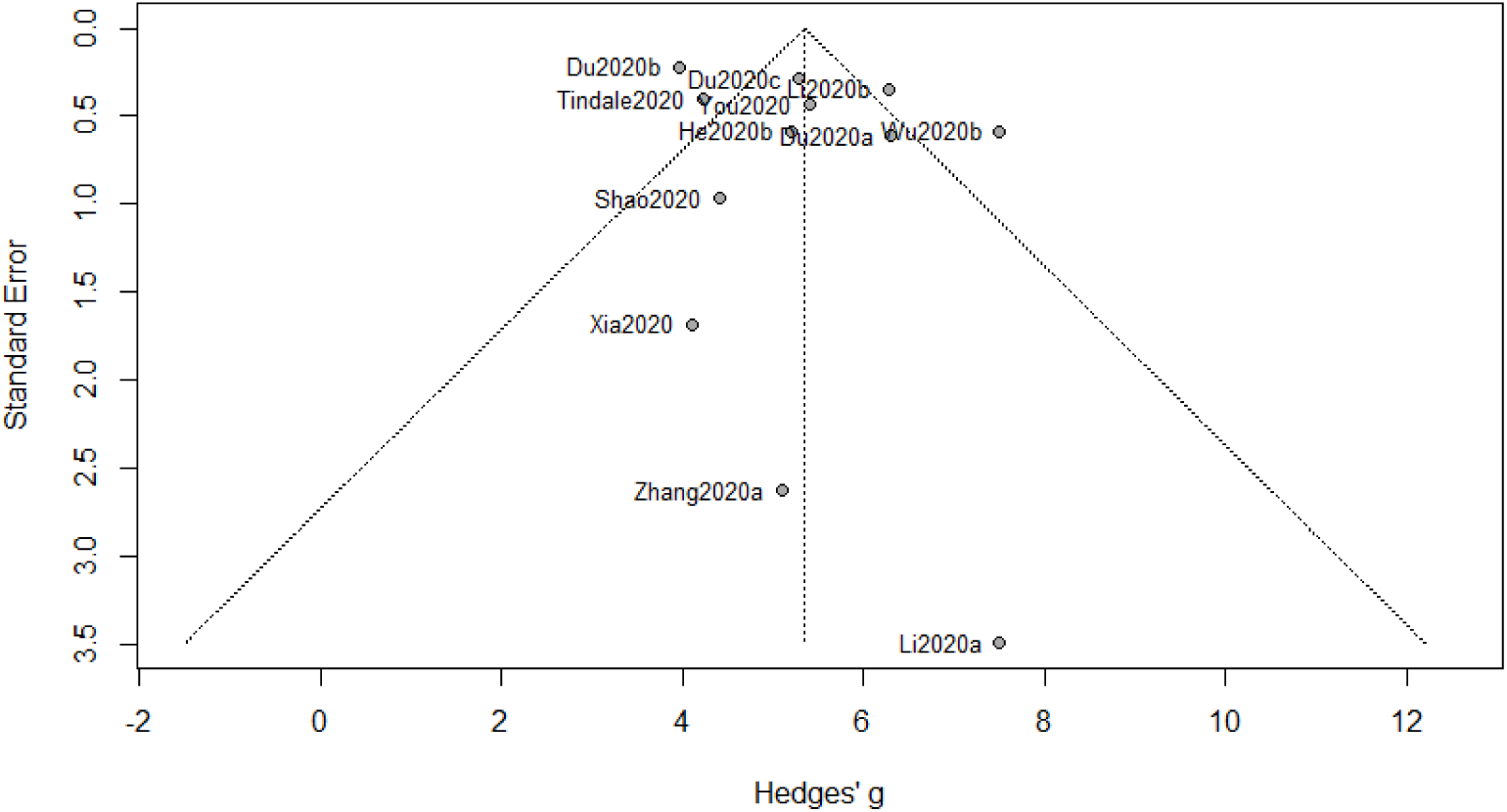
Funnel plot of meta-analysis for serial interval

The estimate of the serial interval of COVID-19 based on our meta-analysis is close to that of SARS (5–6 days according to WHO’s report in “Consensus document on the epidemiology of severe acute respiratory syndrome (SARS)”). A shorter serial interval of COVID-19, together with a shorter mean incubation period, suggests higher possibility that a transmission is completed before the onset of symptoms. Therefore, reducing the source of transmission (by hospitalizing infected individuals or implementing “stay-at-home” protocols to susceptible individuals) and reasonably extending the quarantine period are extensively helpful to slow the progression of COVID-19.

### Epidemic doubling time

The number of the collected studies for epidemic doubling time is 8 (less than 10). As a small sample size is likely to cause bias in meta-analysis (Lin, 2018), the visualization of the funnel plot in Figure 7 exhibits asymmetry.

**Figure 6:**
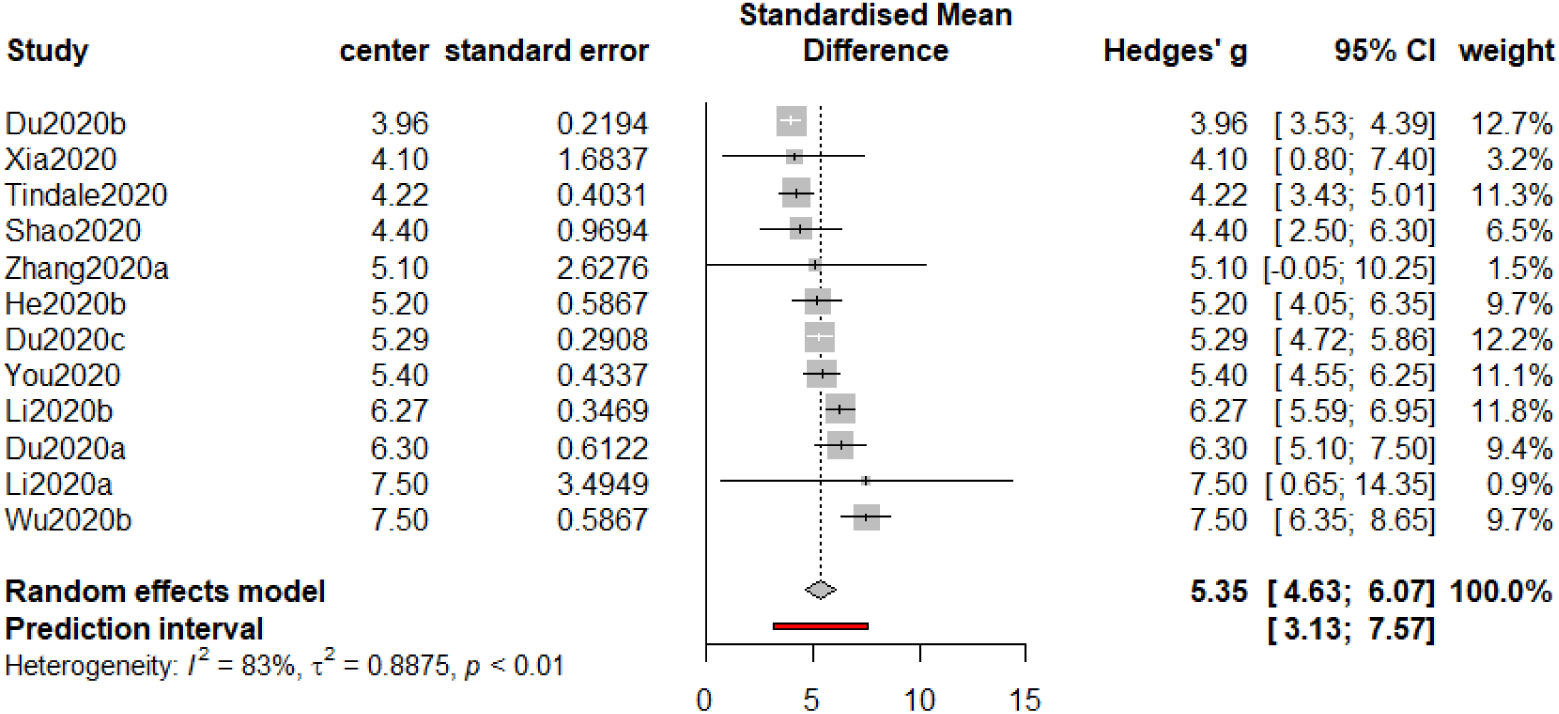
Forest plot of meta-analysis for serial interval

**Figure 7:**
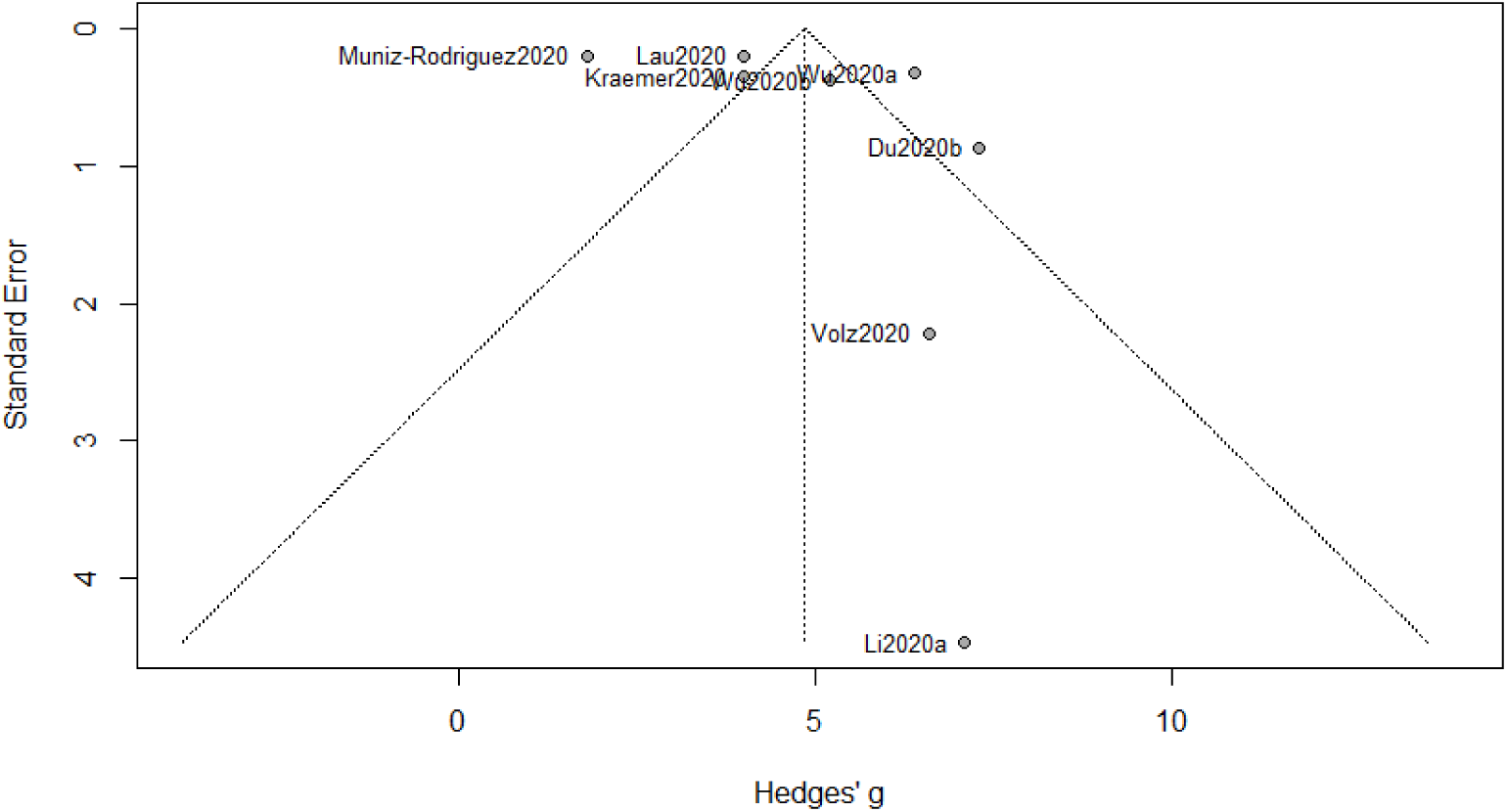
Funnel plot of meta-analysis for epidemic doubling time

We consider the *trim-and-fill procedure* (Duval and Richard, 2000) based on the imputations to reduce the bias owing to a small sample size, and the estimation results before and after implementing the trim-and-fill procedure are given in Figure 8. The estimate without correction is 4.86 (with a 95% confidence interval [3.26, 6.45]), which is larger than the trimmed estimate of value 3.48 (with a 95% confidence interval [1.60, 5.35]). Hence, even though the *p*-value of the Egger’s test is not significant, a remedy is still necessary, as an extensively small sample size usually jeopardizes the statistical power of the Egger’s test. In contrast, when we apply the trim-and-fill procedure to the previous three epidemic metrics, no significant difference has been detected.

**Figure 8:**
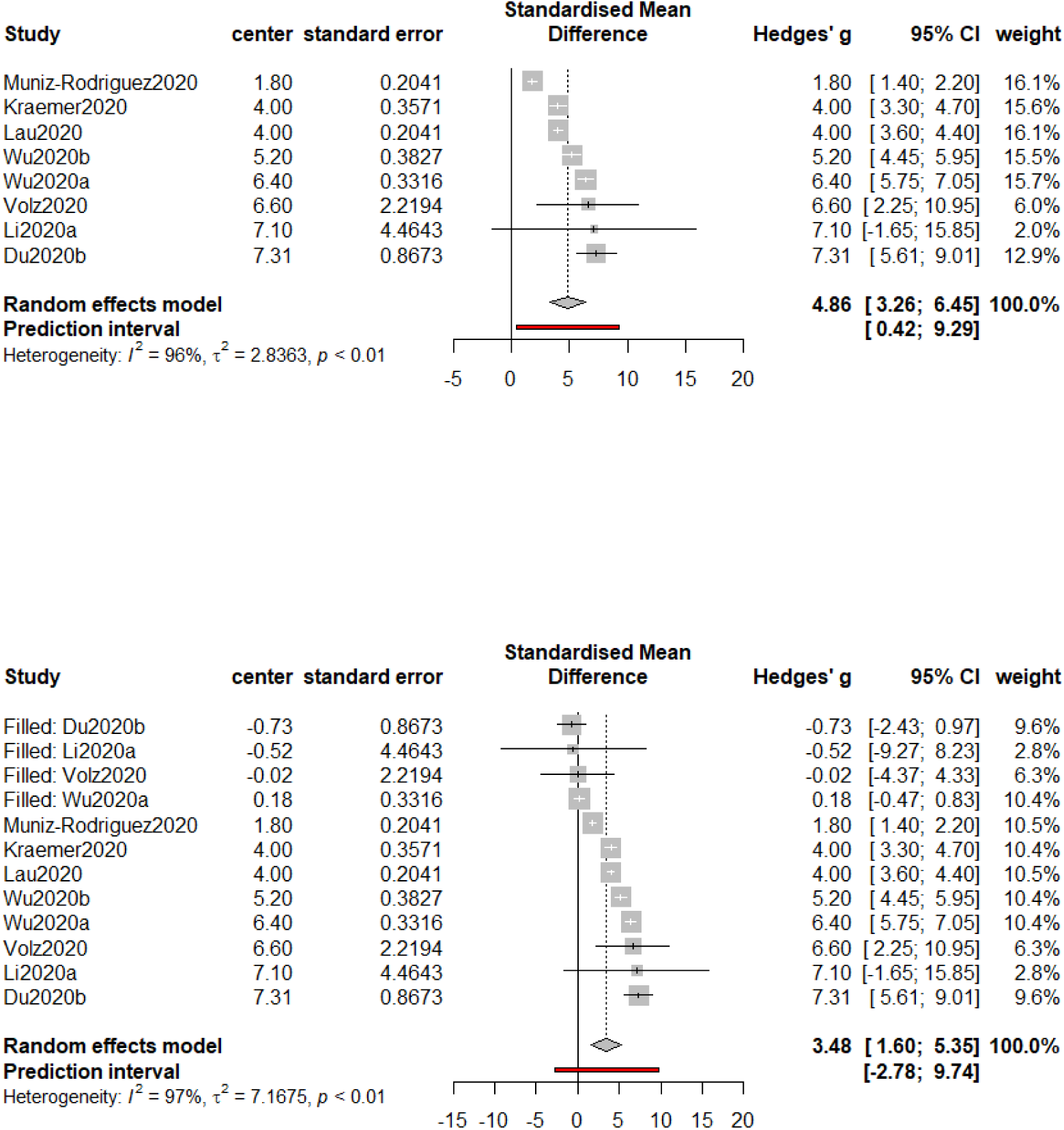
Forest plots of meta-analysis for epidemic doubling time before (top) and after (bottom) implementing the trim-and-fill procedure

Although there seems to be no official report on the epidemic doubling time of SARS from WHO or CDC, we find that an estimate of the doubling time of SARS from a published article (Galvani et al., 2003) is 16.3 through literature search. The estimate of the doubling time of SARS is three times more than the counterpart of COVID-19 based on our analysis, suggesting that COVID-19 is a more contagious disease.

## 6 Sensitivity analysis

Meta-analysis and sensitivity analysis usually go hand in hand. Meta analysis places focus on the summary of a systematic review of relevant studies, whereas the sensitivity analysis is used to assess the robustness of the results from the meta-analysis. In practice, the *sensitivity analysis* is a repeat of the meta-analysis, substituting alternative studies or the results from unclear studies. In other words, the goal of sensitivity analysis is to explore the impact of the meta-analysis by including or excluding studies in the meta-analysis based on some criteria as in Higgins et al. (2019, Section 9.7). In this section, we present a couple of sensitivity analyses for the basic reproduction number *R*_0_. Analogous studies can be carried out for other epidemic characteristics, done mutatis mutandis. For the sake of brevity, we present the details about the sensitivity analysis of *R*_0_ only in this section.

In the first sensitivity analysis, we only include the published articles (a total number of 7 left) in the new analysis, by leaving out preprints that have not yet been through the peer review process. The new estimate of *R*_0_ is 2.80 with a 95% confidence interval [2.05, 3.54]. It is unnecessary to use the prediction interval for inference in the new analysis since we have *I*^2^ = 19%, which is interpreted as unimportant to estimation consistency (Higgins et al., 2019, Section 9.5).

Having observed a small *I*^2^, we conduct a follow-up sensitivity analysis which includes only the 7 published articles and fit a FEM. The estimate of *R*_0_ decreases slightly to 2.70, and the 95% confidence interval gets narrower: [2.62, 2.77].

Now instead of leaving out ambiguous results, we expand the scope of our study to the East Asia. We add several studies from Japan, Korea and the Diamond Princess Cruise, which are listed in Table 6. The estimate of *R*_0_ for this analysis is 2.80 with a 95% confidence interval [2.05, 3.54] and a prediction interval [0.92, 4.69]. With *I*^2^ = 98%, we recommend using the prediction interval for interval-based inference.

## 7 Concluding remarks

In this meta-analysis, we study a collection of recent studies on the COVID-19 pandemic, focusing mainly on four epidemic characteristics: *R*_0_ or the basic reproduction number, the incubation period, the serial interval, and the doubling time of the epidemic. We summarize our numerical findings in Table 1 and include corresponding comparisons with SARS, which is also a viral respiratory illness caused by a coronavirus in 2003. From Table 1, we see that compared to SARS, the COVID-19 has a larger *R*_0_, a longer incubation period and much shorter doubling time, thus suggesting this novel coronavirus be more contagious and stringent public health strategies be necessary.

**Table 1:** Summary of our numerical findings with respect to *R*_0_ or the basic reproduction number, the incubation period, the serial interval and the doubling time of the epidemic.

| Metric | Est. | 95% CI | 95% PI | ≥ or ≤ SARS |
| --- | --- | --- | --- | --- |
| Basic Rep. Num. | 3.16 | [2.60, 3.72] | [1.25, 5.08] | ≥ |
| Incub. Period | 5.35 | [4.29, 6.42] | [1.97, 8.73] | ≥ |
| Serial Int. | 5.35 | [4.63, 6.07] | [3.13, 7.57] | ≈ |
| Epi. Doub. Time | 3.48 | [1.60, 5.35] | $[-2.78, 9.74]$ | ≤ |

The numerical results also provide insights on further studies. For example, with pooled estimates of *R*_0_ and the doubling time available, we can then compare them with the *effective reproduction number* (or *R_t_*) or the doubling time after the nationwide lockdown protocol has been implemented in China to see the effectiveness of the public health strategies. The estimates of the incubation period and the serial interval reassure the necessity of reinforcing a 14-day quarantine period to prevent the spread of the disease.

It is also worthwhile noting the potential limitations of the current meta-analysis. To ensure the accuracy of the estimates collected, we do not include publications that report estimates with a 90% confidence interval or estimates with quartiles. This may lead to some loss of precision or publication bias for our estimates in meta-analysis. Meanwhile, the study on the COVID-19 may not be only restricted to the four characteristics listed in the manuscript. Other metrics, e.g., the case-to fatality rate and the testing capacity, and factors, e.g., significant clusters and patients’ underlying chronic medical conditions, are also of great importance in the future endeavors to mitigate the negative impacts of the COVID-19 pandemic.

## Data Availability

N/A

## Acknowledgments

Dr. Zhang and Dr. Xie received support from the National Institute of Health (NIH) grant R01-NS102324.

## Appendix

In the appendix, we give the details about the studies that are collected for this meta-analysis as well as the graphic representations of the analysis results.

### Basic reproduction number

We list the sources that are utilized for estimating *R*_0_ via meta-analysis in Table 2. In Sun et al. (2020); Zhu and Chen (2020), multiple estimates of *R*_0_ and associated confidence intervals were reported. We select the the most appropriate one based off proposed models, estimation time and other decisive factors.

**Table 2:** Estimates and confidence intervals of *R*_0_ for COVID-19 in China in the literature

| Source | $R_0$ | | |
| --- | --- | --- | --- |
|  | estimate | lower bound | upper bound |
| Cao et al. (2020) | 4.08 | 3.37 | 4.77 |
| Kucharski et al. (2020) | 2.35 | 1.15 | 4.77 |
| Li et al. (2020a) | 2.20 | 1.40 | 3.90 |
| Liu et al. (2020a) | 2.90 | 2.32 | 3.63 |
| Imai et al. (2020) | 2.60 | 1.50 | 3.50 |
| Read et al. (2020) | 3.11 | 2.39 | 4.13 |
| Sanche et al. (2020) | 5.70 | 3.80 | 8.90 |
| Shen et al. (2020) | 4.71 | 4.50 | 4.92 |
| Sun et al. (2020) | 2.94 | 2.38 | 3.50 |
| Tian et al. (2020) | 3.15 | 3.04 | 3.26 |
| Wu et al. (2020a) | 2.68 | 2.47 | 2.86 |
| Zhu and Chen (2020) | 2.69 | 2.61 | 2.77 |

### Incubation period

We list the sources that are utilized for estimating incubation period via meta-analysis in Table 3. In Linton et al. (2020), the authors adopted a variety of distributions for modeling the probability density function of incubation period, leading to slightly different results. We picked the confidence interval under the utilization of log-normal.

**Table 3:** Estimates and confidence intervals of incubation period of COVID-19 in China in the literature incubation period estimate lower bound upper bound

| Source | incubation period |  |  |
| --- | --- | --- | --- |
|  | estimate | lower bound | upper bound |
| Backer et al. (2020) | 6.40 | 5.70 | 7.70 |
| He et al. (2020b) | 2.30 | 0.80 | 3.00 |
| Lauer et al. (2020) | 5.10 | 4.50 | 5.80 |
| Leung (2020) | 6.90 | 5.50 | 8.30 |
| Li et al. (2020a) | 5.20 | 4.10 | 7.00 |
| Linton et al. (2020) | 5.00 | 4.40 | 5.60 |
| Liu et al. (2020a) | 4.80 | 2.20 | 9.40 |
| Men et al. (2020) | 5.84 | 2.91 | 8.75 |
| Qin et al. (2020) | 8.13 | 7.37 | 8.91 |
| Sanche et al. (2020) | 4.20 | 3.50 | 5.10 |
| Xia et al. (2020) | 4.90 | 4.40 | 5.40 |

### Serial interval

We list the sources that are utilized for estimating incubation period via meta-analysis in Table 4. In You et al. (2020), the authors considered three models (maximum likelihood, exponential growth rate and stochastic SIR) and estimated serial intervals for mainland China and a number of provinces. We select the estimate for mainland China via the method of stochastic SIR. In Li et al. (2020b), serial interval estimates of different generations were given, where the estimate for the first generation is chosen for the present analysis.

**Table 4:** Estimates and confidence intervals of serial interval of COVID-19 in China in the literature serial interval estimate 95% lower bound upper bound

| Source | serial interval |  |  |
| --- | --- | --- | --- |
|  | estimate | 95% lower bound | upper bound |
| Du et al. (2020a) | 6.30 | 5.20 | 7.60 |
| Du et al. (2020b) | 3.96 | 3.53 | 4.39 |
| Du et al. (2020c) | 5.29 | 4.72 | 5.86 |
| He et al. (2020b) | 5.20 | 4.10 | 6.40 |
| Li et al. (2020a) | 7.50 | 5.30 | 19.00 |
| Li et al. (2020b) | 6.27 | 5.62 | 6.98 |
| Shao et al. (2020) | 4.40 | 2.90 | 6.70 |
| Tindale et al. (2020) | 4.22 | 3.43 | 5.01 |
| Wu et al. (2020b) | 7.50 | 5.80 | 8.10 |
| Xia et al. (2020) | 4.10 | 0.80 | 7.40 |
| You et al. (2020) | 5.40 | 4.50 | 6.20 |
| Zhang et al. (2020a) | 5.10 | 1.30 | 11.60 |

### Epidemic doubling time

We list the sources that are utilized for estimating epidemic doubling time via meta-analysis in Table 5. In Lau et al. (2020), the authors reported two estimates of epidemic doubling time and their confidence intervals, respectively before and after imposing lockdown in mainland China; we adopt the latter in the analysis. In Muniz-Rodriguez et al. (2020), the authors estimated epidemic doubling times for 31 provinces and municipalities in mainland China; we pick the one for “mainland China (except for Hubei province)” in our study since it is the most comprehensive metric.

**Table 5:** Estimates and confidence intervals of epidemic doubling time of COVID-19 in China in the literature

| Source | epidemic doubling time |  |  |
| --- | --- | --- | --- |
|  | estimate | lower bound | upper bound |
| Du et al. (2020b) | 7.31 | 6.26 | 9.66 |
| Kraemer et al. (2020) | 4.00 | 3.60 | 5.00 |
| Lau et al. (2020) | 4.00 | 3.50 | 4.30 |
| Li et al. (2020a) | 7.10 | 3.00 | 20.50 |
| Muniz-Rodriguez et al. (2020) | 1.80 | 1.50 | 2.30 |
| Volz et al. (2020) | 6.60 | 4.00 | 12.70 |
| Wu et al. (2020a) | 6.40 | 5.80 | 7.10 |
| Wu et al. (2020b) | 5.20 | 4.60 | 6.10 |

### Sensitivity analysis

We list the sources that are utilized for estimating basic reproduction number in sensitivity analysis in Table 6. The added studies include recent research in Japan, South Korea and Diamond Princess Cruise.

**Table 6:** Additional estimates and confidence intervals of basic reproduction number of COVID-19 for sensitivity analysis

| Source | region | $R_0$ | | |
| --- | --- | --- | --- | --- |
|  |  | estimate | lower bound | upper bound |
| Kuniya (2020) | Japan | 2.60 | 2.40 | 2.80 |
| Sugishita et al. (2020) | Japan | 1.99 | 1.89 | 2.09 |
| Shim et al. (2020) | South Korea | 1.50 | 1.40 | 1.60 |
| Zhang et al. (2020b) | Diamond Princess Cruise | 2.28 | 2.06 | 2.52 |
| Zhao et al. (2020) | Diamond Princess Cruise | 2.20 | 2.10 | 2.40 |

## References

Backer, J. A., Klinkenberg, D. and Wallinga, J. (2020). Incubation period of 2019 novel coronavirus (2019-nCoV) infections among travellers from Wuhan, China, 20-28 January 2020. Eurosurveillance, 25, 2000062.

Bi, Q., Wu, Y., Mei, S., Ye, C., Zou, X., Zhang, Z., Liu, X., Wei, L., Truelove, S. A., Zhang, T., Cao, W., Cheng, C., Tang, X., Wu, X., Wu, Y., Sun, B., Huang, S., Sun, Y., Zhang, J., Ma, T., Lessler, J. and Feng, T. (2020). Epidemiology and transmission of COVID-19 in Shenzhen China: Analysis of 391 cases and 1,286 of their close contacts. MedRxiv, https://doi.org/10.1101/2020.03.03.20028423.

Cao, Z., Zhang, Q., Lu, X., Pfeiffer, D., Jia, Z., Song, H. and Zeng, D. D. (2020). *MedRxiv*, https://doi.org/10.1101/2020.01.27.20018952.

Cori, A., Ferguson, N. M., Fraser, C. and Cauchemez, S. (2013). A new framework and software to estimate time-varying reproduction numbers during epidemics. American Journal of Epidemiology, 178, 1505–1512.

Delamater, P. L., Street, E. J., Leslie, T. F., Yang, Y. T. and Jacobsen, K. H. (2019). Complexity of the Basic Reproduction Number (*R*_0_). Emerging Infectious Diseases, 25, 1–4.

Diekmann, O., Heesterbeek, H. and Britton, T. Mathematical Tools for Understanding Infectious Disease Dynamics. Princeton University Press, Princeton, NJ.

Dietz, K. (1993). The estimation of the basic reproduction number for infectious diseases. Statistical Methods in Medical Research, 2, 23–41.

Dong, J., Zhou, Y., Zhang, Y. and Fraz, D. (2020). A validation study for the successful isolation policy in China: A meta-analysis in COVID-19. MedRxiv, https://doi.org/10.1101/2020.04.15.20065102.

Du, Z., Xu, X., Wu, Y., Wang, L. and Cowling, B. J. (2020). Serial interval of COVID-19 from publicly reported confirmed cases. *Emerging Infectious Diseases*, https://doi.org/10.3201/eid2606.200357.

Du, Z., Wang, L., Cauchemez, S., Xu, X., Wang, X., Cowling, B. J. and Meyer, L. A. (2020). Risk for transportation of coronavirus disease from Wuhan to other cities in China. Emerging Infectious Diseases, 26, 1049–1052.

Du, Z., Xu, X., Wu, Y., Wang, L., Cowling, B. J. and Meyers, L. A. (2020). COVID-19 serial interval estimates based on confirmed cases in public reports from 86 Chinese cities. MedRxiv, https://doi.org/10.1101/2020.04.23.20075796.

Duval, S. and Richard, T. (2000). Trim and fill: A simple funnel-plot–based method of testing and adjusting for publication bias in meta-analysis. Biometrics, 56, 455–463.

Egger, M., Davey, S. G., Schneider, M. and Minder, C. (1997). Bias in meta-analysis detected by a simple, graphical test. The BMJ, 315, 629–634.

Fine, P. E. M. (2003). The interval between successive cases of an infectious disease. American Journal of Epidemiology, 158, 1039–1047.

Galvani, A. P., Lei, X. and Jewell, N. P. (2003). Severe acute respiratory syndrome: Temporal stability and geographic variation in death rates and doubling times. Emerging Infectious Diseases, 9, 991–994.

Guan, W.-J., Ni, Z.-Y., Hu, Y., Liang, W.-H., Ou, C.-Q., He, J.-X., Liu, L., Shan, H., Let, C.-L., Hui D. S. C., Du, B., Li, L.-J., Zeng, G., Yuen, K.-Y., Chen, R.-C., Tang, C.-L., Wang, T., Wang, J.-L., Liang, Z.-J., Peng, Y.-X., Wei, L., Liu, Y., Hu, Y.-H., Peng, P., Wang, J.-M., Liu, J.-Y., Chen, Z., Li, G., Zheng, Z.-J., Qiu, S.-Q., Luo, J., Ye, C.-J., Zhu, S.-Y., Zhong, N.-S., and for the China Medical Treatment Expert Group for Covid-19. (2020). Clinical characteristics of coronavirus disease 2019 in China. The New England Journal of Medicine, 382, 1708–1720.

Hartung, J. and Knapp, G. (2001). On tests of the overall treatment effect in meta-analysis with normally distributed responses. Statistics in Medicine, 20, 1771–1780.

He, W., Yi, G. G. and Zhu, Y. (2020). Estimation of the basic reproduction number, average incubation time, asymptomatic infection rate, and case fatality rate for COVID-19: Meta-analysis and sensitivity analysis. MedRxiv, https://doi.org/10.1101/2020.04.28.20083758.

He, X., Lau, E. H. Y., Wu, P., Deng, X., Wang, J., Hao, X., Lau, Y. C., Wong, J. Y., Guan, Y., Tan, X., Mo, X., Chen, Y., Liao, B., Chen, W., Hu, F., Zhang, Q., Zhong, M., Wu, Y., Zhao, L., Zhang, F., Cowling, B. J., Li, F. and Leung, G. M. (2020). Temporal dynamics in viral shedding and transmissibility of COVID-19. Nature Medicine, 26, 672–675.

Higgins, J. P. T., Thomas, J., Chandler, J., Cumpston, M., Li, T., Page, M. J., Welch, V. A. (2019). Cochrane Handbook for Systematic Reviews of Interventions. Second Edition. John Wiley & Sons, Chichester, UK.

Holme, P. and Masuda, N. (2015). The basic reproduction number as a predictor for epidemic outbreaks in temporal networks. PLoS One, 10, e0120567.

Imai, N., Cori, A., Dorigatti, I., Baguelin, M., Donnelly, C. A., Riley, S. and Ferguson, N. M. (2020). Report 3: Transmissibility of 2019-nCoV. Retrieved from https://www.imperial.ac.uk/media/imperial-college/medicine/mrc-gida/2020-01-25-COVID19-Report-3.pdf.

Kraemer, M. U. G., Yang, C.-H., Gutierrez, B., Wu, C.-H., Klein, B., Pigott, D. M., Open COVID-19 Data Working Group, Du Plessis, L., Faria, N. R., Li, R., Hanage, W. P., Brownstein, J. S., Layan, M., Vespignani, A., Tian, H., Dye, C., Pybus, O. G. and Scarpino, S. V. (2020). The effect of human mobility and control measures on the COVID-19 epidemic in China. Nature, 368, 493–497.

Kucharski, A. J., Russell, T. W., Diamond, C., Liu, Y., Edmunds, J., Funk, S. and Eggo, R. M. (2020) Early dynamics of transmission and control of COVID-19: A mathematical modelling study. The Lacent Infectious Diseases, 22, 553–558.

Kuniya, T. (2020). Prediction of the epidemic peak of coronavirus disease in Japan, 2020. Journal of Clinical Medicine, 9, 789.

Lau, H., Khosrawipour, V., Kocbach, P., Mikolajczyk, A., Schubert, J., Bania, J. and Khosrawipour, T. (2020). The positive impact of lockdown in Wuhan on containing the COVID-19 outbreak in China. Journal of Travel Medicine, https://doi.org/10.1093/jtm/taaa037

Lauer, S. A., Grantz, K. H., Bi, Q., Jones, F. K., Zheng, Q., Meredith, H. R., Azman, A. S., Reich, N. G. and Lessler, J. (2020). The incubation period of coronavirus disease 2019 (COVID-19) from publicly reported confirmed cases: Estimation and application. Annals of Internal Medicine, 172, 577–582.

Leung, C. (2020). Estimating the distribution of the incubation period of 2019 novel coronavirus (COVID-19) infection between travelers to Hubei, China and non-travelers. MedRxiv, https://doi.org/10.1101/2020.02.13.20022822.

Li, Q., Guan, X., Wu, P., Wang, X., Zhou, L., Tong, Y., Ren, R., Leung, K. S. M., Lau, E. H. Y., Wong, J. Y., Xing, X., Xiang, N., Wu, Y., Li, C., Chen, Q., Li, D., Liu, T., Zhao, J., Liu, M., Tu, W., Chen, C., Jin, L., Yang, R., Wang, Q., Zhou, S., Wang, R., Liu, H., Luo, Y., Liu, Y., Shao, G., Li, H., Tao, Z., Yang, Y., Deng, Z., Liu, B., Ma, Z., Zhang, Y., Shi, G., Lam, T. T. Y., Wu, J. T., Gao, G. F., Cowling, B. J., Yang, B., Leung. G. M. and Feng, Z. (2020). Early transmission dynamics in Wuhan, China, of novel coronavirus-infected pneumonia. The New England Journal of Medicine, 382, 1199–1207.

Li, M., Liu, K., Song, Y., Wang, M. and Wu, J. (2020). Serial interval and generation interval for respectively the imported and local infectors estimated using reported contact-tracing data of COVID-19 in China. MedRxiv, https://doi.org/10.1101/2020.04.15.20065946.

Lin, L. (2018). Bias caused by sampling error in meta-analysis with small sample sizes. PLoS One, 13, e0204056.

Linton, N. M., Kobayashi, T., Yang, Y., Hayashi, K., Akmetzhanov, A. R., Jung, S.-M., Yuan, B., Kinoshita, R. and Nishiura, H. (2020). Incubation period and other epidemiological characteristics of 2019 novel coronavirus infections with right truncation: A statistical analysis of publicly available case data. Journal of Clinical Medicine, 9, 538.

Lipsitch, M., Cohen, T., Cooper, B., Robins, J. M., Ma, S., James, L., Gopalakrishna, G., Chew, S. K., Tan, C. C., Samore, M. H., Fisman, D. and Murray M. (2020). Transmission dynamics and control of severe acute respiratory syndrom. Science, 300, 1966–1970.

Liu, T., Hu, J., Kang, M., Lin, L., Zhong, H., Xiao, J., He, G., Song, T., Huang, Q., Rong, Z., Deng, A., Zeng, W., Tan, X., Zeng, S., Zhu, Z., Li, J., Wan, D., Lu, J., Deng, H., He, J. and Ma, W. (2020). Transmission dynamics of 2019 novel coronavirus (2019-nCoV). BioRxiv, https://doi.org/10.1101/2020.01.25.919787.

Liu, Y., Gayle, A. A. Wilder-Smith, A. and Rocklöv, J. (2020). The reproductive number of COVID-19 is higher compared to SARS coronavirus. Journal of Travel Medicine, 27, 1–4.

Men, K., Wang, X., Li, Y., Zhang, G., Hu, J., Gao, Y. and Han, H. (2020). Estimate the incubation period of coronavirus 2019 (COVID-19). MedRxiv, https://doi.org/10.1101/2020.02.24.20027474.

Muniz-Rodriguez, K., Chowell, G., Cheung, C.-H., Jia, D., Lai, P.-Y., Lee, Y., Liu, M., Ofori, S. K., Roosa, K. M., Simonsen, L., Viboud, C. and Fung, I. C.-H. (2020). Doubling time of the COVID-19 epidemic by Chinese province. MedRxiv, https://doi.org/10.1101/2020.02.05.20020750.

Nikbakht, R., Baneshi, M. R., Bahrampour, A. and Hosseinnataj, A. (2019). Comparison of methods to estimate basic reproduction number (*R* − 0) of influenza, using Canada 2009 and 2017-18 A (H1N1) data. Journal of Research in Medical Sciences, 24, 67.

Qin, J., You, C., Lin, Q., Hu, T., Yu, S. and Zhou, X.-H. (2020). Estimation of incubation period distribution of COVID-19 using disease onset forward time: A novel cross-sectional and forward follow-up study. MedRxiv, https://doi.org/10.1101/2020.03.06.20032417

Read, J. M., Bridgen, J. R. E., Cummings, D. A. T., Ho, A. and Jewell, C. P. (2020). Novel coronavirus 2019-nCoV: early estimation of epidemiological parameters and epidemic predictions. MedRxiv, https://doi.org/10.1101/2020.01.23.20018549.

Reich, N. G., Lessler, J., Cummings, D. A. and Brookmeyer, R. (2009). Estimating incubation period distributions with coarse data. Statistics in Medicine, 28, 2769–2784.

Sanche, S., Lin, Y. T., Xu, C., Romero-Severson, E., Hengartner, N. and Ke, R. (2020). High contagiousness and rapid spread of severe acute respiratory syndrome coronavirus 2. Emerging Infectious Diseases. https://doi.org/10.3201/eid2607.200282.

Shao, S., Gao, D., Zhuang, Z., Chong, M. K. C., Cai, Y., Ran, J., Cao, P., Wang, K., Lou, Y., Wang, W., Yang, L., He, D. and Wang, M. H. (2020). Estimating the serial interval of the novel coronavirus disease (COVID-19): A statistical analysis using the public data in Hong Kong from January 16 to February 15, 2020. MedRxiv, https://doi.org/10.1101/2020.02.21.20026559

Shen, M., Peng, Z., Xiao, Y. and Zhang, L. (2020). Modelling the epidemic trend of the 2019 novel coronavirus outbreak in China. BioRxiv, https://doi.org/10.1101/2020.01.23.916726.

Shim, E., Tariq, A., Choi, W., Lee, Y. and Chowell, G. (2020). Transmission potential and severity of COVID-19 in South Korea. International Journal of Infectious Diseases, 93, 339–344.

Sidik, K. and Jonkman, J. N. (2002). A simple confidence interval for meta-analysis. Statistics in Medicine, 21, 3153–3159.

Sugishita, Y., Kurita, J., Sugawara, T. and Ohkusa, Y. (2020). Effect of voluntary event cancellation and school closure as countermeasures against COVID-19 outbreak in Japan. MedRxiv, https://doi.org/10.1101/2020.03.19.20037945.

Sun, H., Qiu, Y., Yan, H., Huang, Y., Zhu, Y. and Chen, S. (2020). Tracking and predicting COVID-19 epidemic in China mainland. BioRxiv, https://doi.org/10.1101/2020.02.17.20024257.

Tian, H., Liu, Y., Li, Y., Wu, C.-H., Chen, B., Kraemer, M. U. G., Li, B., Cai, J., Xu, B., Yang, Q., Wang, B., Yang, P., Cui, Y., Song, Y., Zheng, P., Wang, Q., Bjornstad, O. N., Yang, R., Grenfell, B., Pybus, O. G. and Dye, C. (2020). An investigation of transmission control measures during the first 50 days of the COVID-19 epidemic in China. Science, 368, 638–642.

Tindale, L. C., Coobe, M., Stockdale, J. E., Garlock, E. S., Lau, W. Y. V., Saraswat, M., Brian, Y.-H., Zhang, L., Chen, D., Wallinga, J. and Colijn, C. (2020). Transmission interval estimates suggest pre-symptomatic spread of COVID-19. MedRxiv, https://doi.org/10.1101/2020.03.03.20029983.

Veroniki, A. A., Jackson, D., Viechtbauer, W., Bender, R., Bowden, J., Knapp, G., Kuss, O., Higgins, J. P. T., Langan, D. and Salanti, G. (2016). Methods to estimate the between-study variance and its uncertainty in meta-analysis. Research Synthesis Methods, 7, 55–79.

Volz, E., Baguelin, M., Bhatia, S., Boonyasiri, A., Cori, A., Cucunubá, Z., Cuomo-Dannenburg, G., Donnelly, C. A., Dorigatti, I., FitzJohn, R., Fu, H., Gaythorpe, K., Ghani, A., Hamlet, A., Hinsley, W., Imai, N., Laydon, D., Nedjati-Gilani, G., Okell, L., Riley, S. Van Elsland, S., Wang, H., Wang, Y., Xi, X. and Ferguson, N. M. (2020). Report 5: Phylogenetic analysis of SARS-CoV-2. Retrieved from https://www.imperial.ac.uk/media/imperial-college/medicine/sph/ide/gida-fellowships/Imperial-College-COVID19-phylogenetics-15-02-2020.pdf.

White, L. F., Wallinga, J., Finelli, L., Reed, C., Riley, S., Lipsitch, M. and Pagano, M. (2009). Estimation of the reproductive number and the serial interval in early phase of the 2009 Influenza A/H1N1 pandemic in the USA. Influenza and Other Respiratory Viruses, 3, 267–276.

Worldometers. Reported cases and deaths by country, territory, or conveyance. https://www.worldometers.info/coronavirus, 2020. Accessed: 2020-05-09.

Wu, J. T., Leung, K. and Leung, G. M. (2020). Nowcasting and forecasting the potential domestic and international spread of the 2019-nCoV outbreak originating in Wuhan, China: A modelling study. The Lancet, 395, 689–697.

Wu, J. T., Leung, K., Bushman, M., Kishore, N., Niehus, R., De Salazar, P. M., Cowling, B. J., Lipsitch, M. and Leung, G. M. (2020). Estimating clinical severity of COVID-19 from the transmission dynamics in Wuhan, China. Nature Medicine, 26, 506–510.

Xia, W., Liao, J., Li, C., Li, Y., Qian, X., Sun, X., Xu, H., Mahai, G., Zhao, X., Shi, L., Liu, J., Yu, L., Wang, M., Wang, Q., Namat, A., Li, Y., Qu, J., Liu, Q., Lin, X., Cao, S., Huan, S., Xiao, J., Ruan, F., Wang, H., Xu, Q., Ding X., Fang, X., Qiu, F., Ma, J., Zhang, Y., Wang, A., Xing, Y. and Xu, S. (2020). Transmission of corona virus disease 2019 during the incubation period may lead to a quarantine loophole. MedRxiv, https://doi.org/10.1101/2020.03.06.20031955.

You, C., Deng, Y., Hu, W., Sun, J., Lin, Q., Zhou, F., Pang C. H., Zhang, Y., Chen, Z. and Zhou, X.-H. (2020). Estimation of the time-varying reproduction number of COVID-19 outbreak in China. MedRxiv, https://doi.org/10.1101/2020.02.08.20021253.

Zhang, J., Litvinova, M., Wang, W., Wang, Y., Deng, X., Chen, X., Li, M., Zheng, W., Yi, L., Chen, X., Wu, Q., Liang, Y., Wang, X., Yang, J., Sun, K., Longini, I. M. Jr., Halloran, M. E., Wu, P., Cowling, B. J., Merler, S., Viboud, C., Vespignani, A., Ajelli, M. and Yu, H. (2020). Evolving epidemiology and transmission dynamics of coronavirus disease 2019 outside Hubei province, China: A descriptive and modelling study. The Lancet Infectious Diseases, https://doi.org/10.1016/S1473-3099(20)30230-9.

Zhang, S., Diao, M., Yu, W., Pei, L., Lin, Z. and Chen, D. Estimation of the reproductive number of novel coronavirus (COVID-19) and the probable outbreak size on the Diamond Princess cruise ship: A data-driven analysis. International Journal of Infectious Diseases, 93, 201–204.

Zhao, S., Gao, P., Gao, D., Zhuang, Z., Chong, M. K. C., Cai, Y., Ran J., Wang, K., Lou, Y., Wang, M., Yang, L., He, D. and Wang, M. H. (2020). Modelling the coronavirus disease (COVID-19) outbreak on the Diamond Princess ship using the public surveillance data from January 20 to February 20, 2020. MedRxiv, https://doi.org/10.1101/2020.02.26.20028449.

Zhu, Y. and Chen, Y.-Q. (2020). On a statistical transmission model in analysis of the early phase of COVID-19 outbreak. Statistics in Biosciences, https://doi.org/10.1007/s12561-020-09277-0.

